# Association of cognitive impairment with adverse childhood experiences: 2019 Results from 21 states

**DOI:** 10.1101/2025.11.05.25339606

**Authors:** Mary L. Adams, Joseph Grandpre

## Abstract

Our objective was to assess associations between 8 adverse childhood experiences (ACEs; physical, emotional, and sexual abuse as a child, or living in household with substance abuse, mental health problems, divorce, intimate partner violence, or incarceration) and subjective cognitive impairment (SCI: the disability question from the 2019 Behavioral Risk Factor Surveillance Surveys) and compare with cardiovascular disease (CVD) results.

**Methods:** Adults (N=149,801) from 21 states with data on ACEs were studied using Stata in unadjusted and adjusted analysis controlled for demographics, selected health measures, CVD, and 6 CVD risk factors (smoking, overweight, hypertension, high cholesterol, diabetes, and inadequate physical activity).

**Results:** Mean number of ACEs ranged from 0.8 among adults age 75+ to 2.2 for adults 18-34 years and 3.0 for adults with SCI. Prevalence rates were 12.6% for SCI, increasing with both number of ACES and CVD risk factors and 9.4% for CVD, only increasing with more CVD risk factors. Among age groups, SCI was highest for adults 18-24 years (17.2%) and lowest for those 65-74 years 9.4%) while CVD was highest among adults age 75+ (26.5%) and lowest for those 18-24 years (1.1%). Most results were confirmed by logistic regression with adjusted odds ratios (AOR) for SCI for each added ACE = 1.20 (95% CI 1.17-1.23) and each added CVD risk factor =1.10 (1.06-1.14) and for CVD AOR= 1.06 (1.03-1.08) for each added ACE and 1.42 (1.38-1.46) for each added risk factor.

**Conclusion:** Results suggest that ACEs may be a factor contributing to higher rates of SCI among younger vs. older adults.

## Introduction

Adverse childhood experiences (ACEs) have been found to be associated with a wide range of poor health outcomes.^1^ These ACEs include physical, emotional, and sexual abuse of the child, plus anyone in the household with substance abuse or mental health problems, divorce, intimate partner violence, or incarceration.^1^ Poor health outcomes include heart disease, cancer, lung and liver disease,^2^ and depression.^3–5^ Evidence from more recent studies ^6–8^ suggests that ACEs may also be associated with cognitive issues in later life. ACEs may impact the parts of the brain that affect reasoning and problem solving (fluid cognition) and crystalized cognition (knowledge acquired over lifetime).^8^ These recent studies on cognition ^6–8^ are promising but limited in the number of participants.

Cognitive impairment or difficulties may be an early step in the development of dementia and Alzheimer’s Disease which is a leading cause of death.^9^ A measure of cognitive impairment is currently required on federal surveys ^10,11^ such as the Behavioral Risk Factor Surveillance System (BRFSS) which is a large, representative telephone survey addressing a wide range of health measures.^12^ A recent paper using BRFSS data found that between 2013 and 2023 the cognitive impairment measure among adults 18-39 years nearly doubled while rates for other age groups showed modest increases or even a decrease.^13^ In addition to cognitive impairment, the BRFSS has addressed ACEs in different years.^1,4^ Our goal for this current study was to study associations between ACEs and this subjective cognitive impairment (SCI) measure to assess the possibility that ACEs may have a role in this increase in prevalence of SCI among younger adults. To add some perspective, we also examine cardiovascular disease (CVD, including heart disease and stroke) and its potentially modifiable risk factors which have been extensively studied.^14,15^ often in comparison with cognitive measures.

## Methods

### Data

We used publicly available 2019 Behavioral Risk Factor Surveillance System (BRFSS) data ^12^ from 149,801 adults ages 18 and older in the 21 states (AL, DE, FL, IN, IA, MI, MS, MO, NM, ND, PA, RI, SC, TN, VA, WV, WI, OH, KS, OK, and NY) that asked the optional ACEs module. Data were adjusted for the probability of selection and weighted to be representative of the adult population in each state by age, gender, race/ethnicity, marital status, education, home ownership, and telephone type. Data sets included weights and stratum variables needed for analysis. Data for states using multiple questionnaire versions were downloaded separately and combined according to BRFSS module protocol.^16^ The median response rate for cell phone and land line surveys combined for the 21 states was 49.6%, ranging from 37.3% in New York to 60.8% in North Dakota. ^17^

### Measures

Subjective cognitive impairment (SCI) was defined as a “yes” response to “Because of a physical, mental, or emotional problem, do you have difficulty remembering, concentrating, or making decisions?” This disability question has been asked since 2008 by the Census Bureau but the measure should not be considered cognitive decline because the question lacks a time frame.^18,19^ ACEs measures from the BRFSS^1^ included separate measures of physical, emotional, and sexual abuse as a child, or living in household with substance abuse, mental health problems, divorce, intimate partner violence (IPV), or incarceration, Also included were composite measures for respondents with valid responses to all 8 questions while excluding 33,769 respondents with missing values. Cardiovascular disease (CVD) included those ever told they had a heart attack, angina, coronary heart disease, or stroke. CVD risk factors included smoking, hypertension, high cholesterol, diabetes, not meeting recommendations for moderate physical activity and having body mass index ≥25 based on self-reported height and weight. Because the BRFSS is a cross-sectional survey, ever smoking was used in lieu of current smoking; ever smokers were respondents who smoked 100 cigarettes in their lifetime. A composite measure that included all 6 CVD risk factors was used in analysis while 28,246 respondents with missing values for any risk factor were excluded.

### Other variables

Demographic measures included gender, age (18-24, 25-34, 35-44, 45-54, 55-64, 65-74, and 75 years and older), self-reported race/ethnicity (non-Hispanic white, Black or African American, Hispanic of any race, American Indian/Alaska Native, Asian, and other), education (college graduate, some college, high school graduate, < high school), household income (≥$75,000, $50,000-$74,999, $25,000-$49,999, $15,000-24,999, <$15,000, and unknown). Depression included anyone ever told they had a depressive disorder, (including depression, major depression, dysthymia or minor depression). Health status was dichotomized as self-reported fair or poor as opposed to good, very good, or excellent. Difficulty seeing was one of the 6 disability questions: “Are you blind or do you have serious difficulty seeing, even when wearing glasses?”

### Statistical Analysis

Stata version 18.0 (Stata Corp LP, College Station, TX) was used for analysis to account for the complex sample design of the BRFSS and used weights and stratum variables supplied in the data set. Data were weighted to account for the probability of selection and further adjusted through a “raking” process to represent the adult population in each state by age, gender, race/ethnicity, marital status, education, home ownership, and telephone source. Point estimates and 95% confidence intervals were determined for SCI and CVD and by demographics, depression, vision problems, and health status, the number of ACEs (0-8) and CVD risk factors (0-6). Mean numbers of ACEs were also determined. In adjusted analysis, the other outcome (e.g. SCI or CVD and composite measures of ACEs and CVD risk factors were added to the model. For both ACEs and CVD risk factors, composite measures were entered as either a continuous variable where each unit change was measured or as a step-wise measure with each number of components (1-8 or 1-6 respectively) being compared with those with 0 ACEs or risk factors, to ascertain dose-response gradients. When entered as a continuous variable, the result represents the adjusted odds ratio for each additional ACE or CVD risk factor.

## Results

The mean number of ACEs was 1.71 (95% CI 1.69-1.73) overall, ranging from 0.81 among adults age 75+ years and 1.04 (1.01-1.06) for ages 65+ (not shown) to 2.19 and 2.21 for those ages 18-24 and 25-34 years respectively (Table 1), with highest mean number of ACEs among groups studied being 2.96 for adults with SCI, and 2.85 for those with depression. About two-thirds (64.5%) of adults reported at least one ACE, 17.8% reported 4 or more, and 0.7% reported all 8 (not shown). For CVD risk factors, 92.5% of adults reported any of the 6 risk factors and 21.6% reported 4 or more.

**Table 1.**
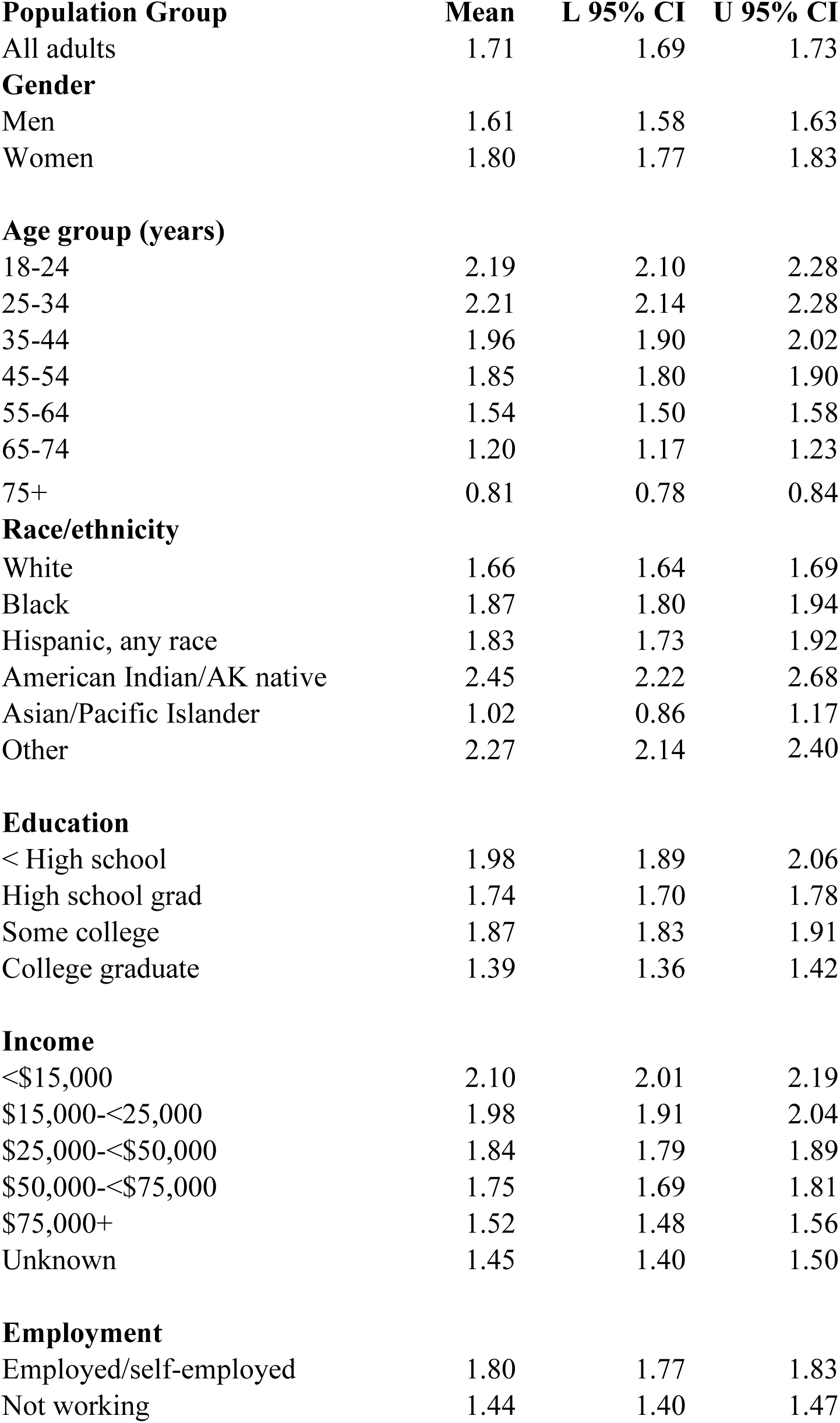
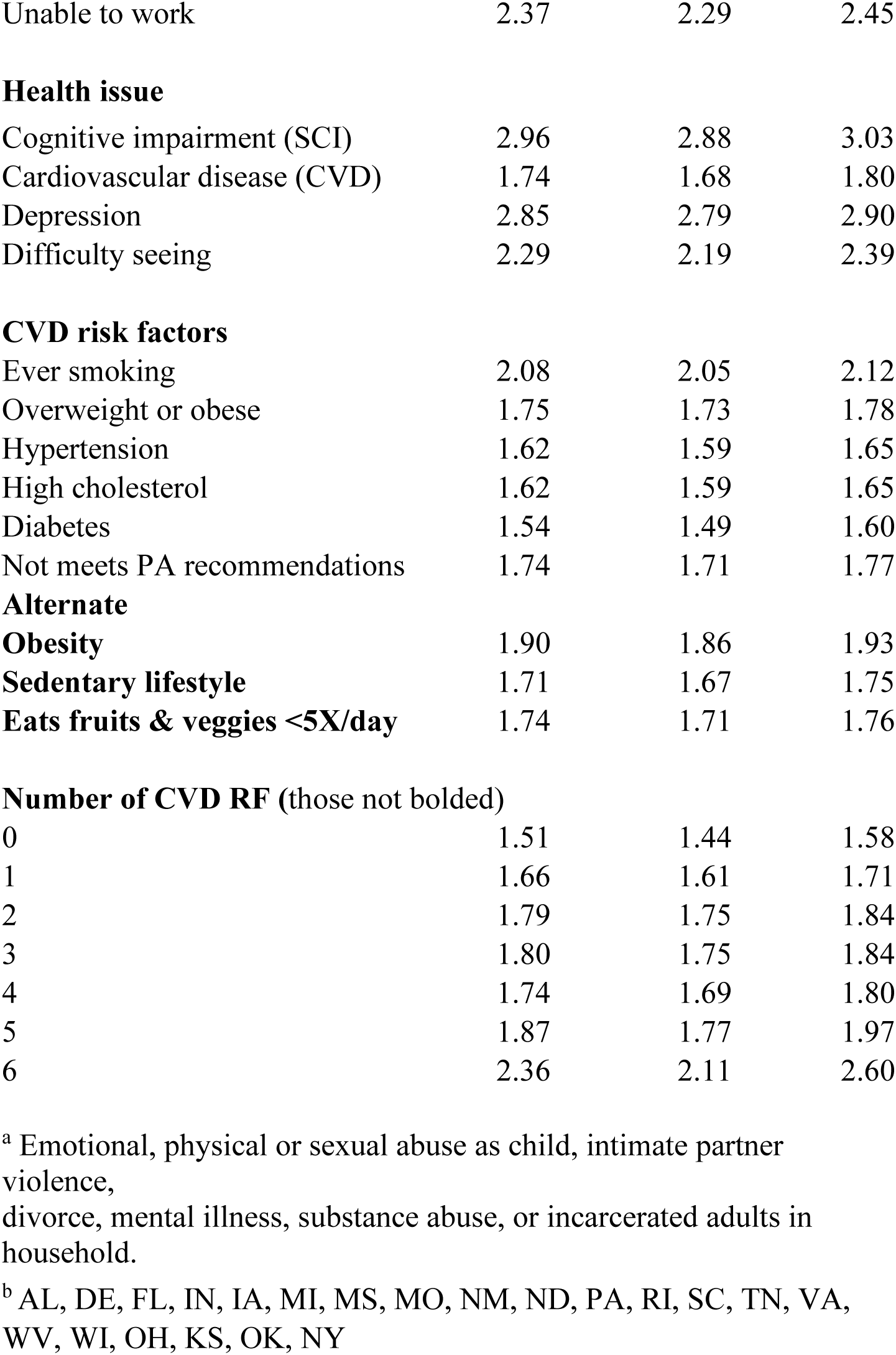
Mean number of adverse childhood experiences ^a^ (ACEs); 2019 Behavioral Risk Factor Surveillance System (BRFSS), 21 states ^b^;

Prevalence rates for SCI and CVD are reported in Table 2 by demographics and other measures with overall prevalence rates of 12.6 and 9.4% respectively. SCI rates ranged from 5.0% for adults with income >$75K to 44.1% for adults with all 8 ACEs. CVD rates ranged from 1.0% for 0 CVD risk factors or 1.1% for 18–24-year-olds to 42.5% for adults with all 6 CVD risk factors. Results for SCI by age ranged from 9.4% among adults ages 65-74 years to 17.2% among those ages 18-24 while CVD rates consistently increased with increasing age. Results for SCI and CVD were generally similar for race, education, income, health status, vision, depression, and association with CVD risk factors, but only SCI was associated with increasing number of ACEs in unadjusted results. Prevalence rates of the separate ACEs are shown in Table 3 for all 21 states, along with results for age groups.

**Table 2.**
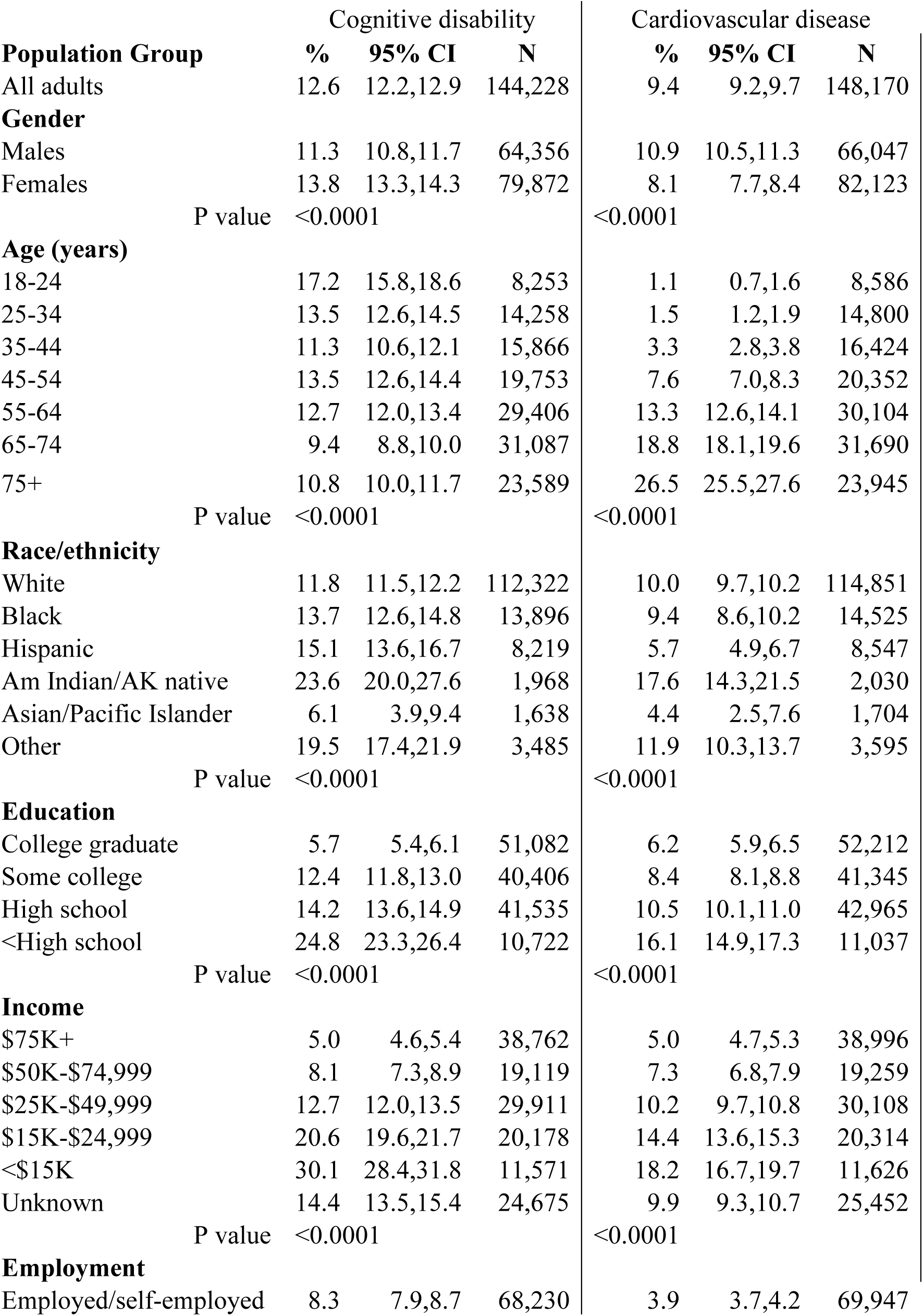
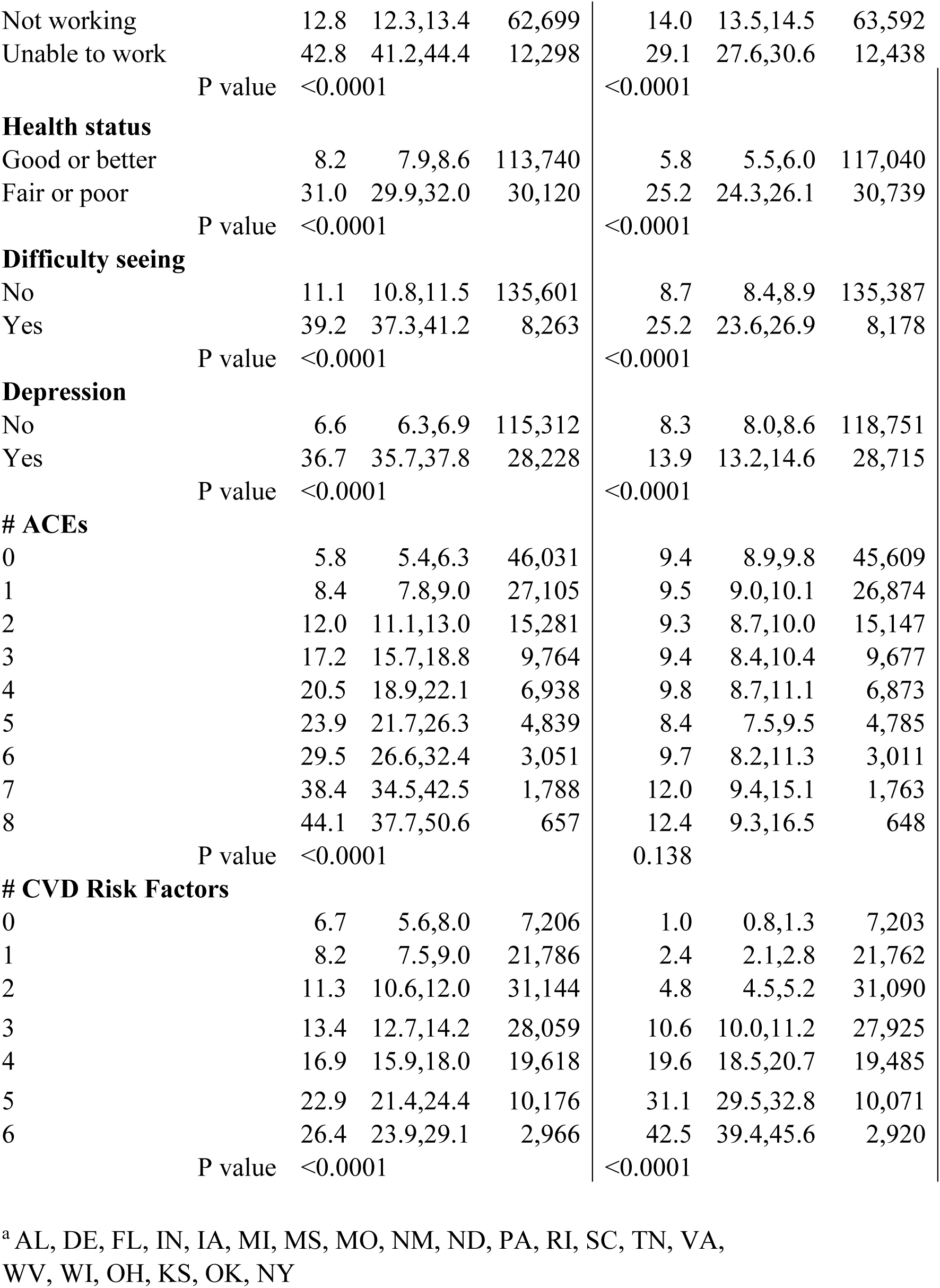
Rates of cognitive impairment and cardiovascular disease by demographic and other measures, 2019 Behavioral Risk Factor Surveillance System (BRFSS), 21 states ^a^.

| **Population Group** | **Cognitive disability** |  |  | **Cardiovascular disease** |  |  |
| --- | --- | --- | --- | --- | --- | --- |
|  | **%** | **95% CI** | **N** | **%** | **95% CI** | **N** |
| All adults | 12.6 | 12.2,12.9 | 144,228 | 9.4 | 9.2,9.7 | 148,170 |
| **Gender** |  |  |  |  |  |  |
| Males | 11.3 | 10.8,11.7 | 64,356 | 10.9 | 10.5,11.3 | 66,047 |
| Females | 13.8 | 13.3,14.3 | 79,872 | 8.1 | 7.7,8.4 | 82,123 |
| P value | <0.0001 |  |  | <0.0001 |  |  |
| **Age (years)** |  |  |  |  |  |  |
| 18-24 | 17.2 | 15.8,18.6 | 8,253 | 1.1 | 0.7,1.6 | 8,586 |
| 25-34 | 13.5 | 12.6,14.5 | 14,258 | 1.5 | 1.2,1.9 | 14,800 |
| 35-44 | 11.3 | 10.6,12.1 | 15,866 | 3.3 | 2.8,3.8 | 16,424 |
| 45-54 | 13.5 | 12.6,14.4 | 19,753 | 7.6 | 7.0,8.3 | 20,352 |
| 55-64 | 12.7 | 12.0,13.4 | 29,406 | 13.3 | 12.6,14.1 | 30,104 |
| 65-74 | 9.4 | 8.8,10.0 | 31,087 | 18.8 | 18.1,19.6 | 31,690 |
| 75+ | 10.8 | 10.0,11.7 | 23,589 | 26.5 | 25.5,27.6 | 23,945 |
| P value | <0.0001 |  |  | <0.0001 |  |  |
| **Race/ethnicity** |  |  |  |  |  |  |
| White | 11.8 | 11.5,12.2 | 112,322 | 10.0 | 9.7,10.2 | 114,851 |
| Black | 13.7 | 12.6,14.8 | 13,896 | 9.4 | 8.6,10.2 | 14,525 |
| Hispanic | 15.1 | 13.6,16.7 | 8,219 | 5.7 | 4.9,6.7 | 8,547 |
| Am Indian/AK native | 23.6 | 20.0,27.6 | 1,968 | 17.6 | 14.3,21.5 | 2,030 |
| Asian/Pacific Islander | 6.1 | 3.9,9.4 | 1,638 | 4.4 | 2.5,7.6 | 1,704 |
| Other | 19.5 | 17.4,21.9 | 3,485 | 11.9 | 10.3,13.7 | 3,595 |
| P value | <0.0001 |  |  | <0.0001 |  |  |
| **Education** |  |  |  |  |  |  |
| College graduate | 5.7 | 5.4,6.1 | 51,082 | 6.2 | 5.9,6.5 | 52,212 |
| Some college | 12.4 | 11.8,13.0 | 40,406 | 8.4 | 8.1,8.8 | 41,345 |
| High school | 14.2 | 13.6,14.9 | 41,535 | 10.5 | 10.1,11.0 | 42,965 |
| <High school | 24.8 | 23.3,26.4 | 10,722 | 16.1 | 14.9,17.3 | 11,037 |
| P value | <0.0001 |  |  | <0.0001 |  |  |
| **Income** |  |  |  |  |  |  |
| \$75K+ | 5.0 | 4.6,5.4 | 38,762 | 5.0 | 4.7,5.3 | 38,996 |
| \$50K-\$74,999 | 8.1 | 7.3,8.9 | 19,119 | 7.3 | 6.8,7.9 | 19,259 |
| \$25K-\$49,999 | 12.7 | 12.0,13.5 | 29,911 | 10.2 | 9.7,10.8 | 30,108 |
| \$15K-\$24,999 | 20.6 | 19.6,21.7 | 20,178 | 14.4 | 13.6,15.3 | 20,314 |
| <\$15K | 30.1 | 28.4,31.8 | 11,571 | 18.2 | 16.7,19.7 | 11,626 |
| Unknown | 14.4 | 13.5,15.4 | 24,675 | 9.9 | 9.3,10.7 | 25,452 |
| P value | <0.0001 |  |  | <0.0001 |  |  |
| **Employment** |  |  |  |  |  |  |
| Employed/self-employed | 8.3 | 7.9,8.7 | 68,230 | 3.9 | 3.7,4.2 | 69,947 |
| Not working | 12.8 | 12.3,13.4 | 62,699 | 14.0 | 13.5,14.5 | 63,592 |
| Unable to work | 42.8 | 41.2,44.4 | 12,298 | 29.1 | 27.6,30.6 | 12,438 |
| P value | <0.0001 |  |  | <0.0001 |  |  |
| Health status |  |  |  |  |  |  |
| Good or better | 8.2 | 7.9,8.6 | 113,740 | 5.8 | 5.5,6.0 | 117,040 |
| Fair or poor | 31.0 | 29.9,32.0 | 30,120 | 25.2 | 24.3,26.1 | 30,739 |
| P value | <0.0001 |  |  | <0.0001 |  |  |
| Difficulty seeing |  |  |  |  |  |  |
| No | 11.1 | 10.8,11.5 | 135,601 | 8.7 | 8.4,8.9 | 135,387 |
| Yes | 39.2 | 37.3,41.2 | 8,263 | 25.2 | 23.6,26.9 | 8,178 |
| P value | <0.0001 |  |  | <0.0001 |  |  |
| Depression |  |  |  |  |  |  |
| No | 6.6 | 6.3,6.9 | 115,312 | 8.3 | 8.0,8.6 | 118,751 |
| Yes | 36.7 | 35.7,37.8 | 28,228 | 13.9 | 13.2,14.6 | 28,715 |
| P value | <0.0001 |  |  | <0.0001 |  |  |
| # ACEs |  |  |  |  |  |  |
| 0 | 5.8 | 5.4,6.3 | 46,031 | 9.4 | 8.9,9.8 | 45,609 |
| 1 | 8.4 | 7.8,9.0 | 27,105 | 9.5 | 9.0,10.1 | 26,874 |
| 2 | 12.0 | 11.1,13.0 | 15,281 | 9.3 | 8.7,10.0 | 15,147 |
| 3 | 17.2 | 15.7,18.8 | 9,764 | 9.4 | 8.4,10.4 | 9,677 |
| 4 | 20.5 | 18.9,22.1 | 6,938 | 9.8 | 8.7,11.1 | 6,873 |
| 5 | 23.9 | 21.7,26.3 | 4,839 | 8.4 | 7.5,9.5 | 4,785 |
| 6 | 29.5 | 26.6,32.4 | 3,051 | 9.7 | 8.2,11.3 | 3,011 |
| 7 | 38.4 | 34.5,42.5 | 1,788 | 12.0 | 9.4,15.1 | 1,763 |
| 8 | 44.1 | 37.7,50.6 | 657 | 12.4 | 9.3,16.5 | 648 |
| P value | <0.0001 |  |  | 0.138 |  |  |
| # CVD Risk Factors |  |  |  |  |  |  |
| 0 | 6.7 | 5.6,8.0 | 7,206 | 1.0 | 0.8,1.3 | 7,203 |
| 1 | 8.2 | 7.5,9.0 | 21,786 | 2.4 | 2.1,2.8 | 21,762 |
| 2 | 11.3 | 10.6,12.0 | 31,144 | 4.8 | 4.5,5.2 | 31,090 |
| 3 | 13.4 | 12.7,14.2 | 28,059 | 10.6 | 10.0,11.2 | 27,925 |
| 4 | 16.9 | 15.9,18.0 | 19,618 | 19.6 | 18.5,20.7 | 19,485 |
| 5 | 22.9 | 21.4,24.4 | 10,176 | 31.1 | 29.5,32.8 | 10,071 |
| 6 | 26.4 | 23.9,29.1 | 2,966 | 42.5 | 39.4,45.6 | 2,920 |
| P value | <0.0001 |  |  | <0.0001 |  |  |

<sup>a</sup> AL, DE, FL, IN, IA, MI, MS, MO, NM, ND, PA, RI, SC, TN, VA, WV, WI, OH, KS, OK, NY

**Table 3.** Separate adverse childhood experiences (ACEs) 2019 BRFSS, 21 states^a^; N=114,165.

| Separate ACEs | All ages<br>N=114,165) |  | Age 18-24<br>N=6,239 |  | Age 75+<br>19,283 |  |
| --- | --- | --- | --- | --- | --- | --- |
| Number in age group>>> | 114,165 |  | 6,239 |  | 19,283 |  |
| **Abuse** | **%** | **95% CI** | **%** | **95% CI** | **%** | **95% CI** |
| Emotional abuse | 34.1 | 33.6-34.7 | 45.8 | 43.7-47.9 | 14.8 | 13.9-15.8 |
| Physical abuse | 24.2 | 23.7-24.7 | 25.3 | 23.5-27.1 | 16.9 | 15.9-17.9 |
| Sexual abuse | 12.8 | 12.5-13.2 | 12.3 | 11.0-13.7 | 7.1 | 6.5-7.8 |
| **Household Challenges** |  |  |  |  |  |  |
| Intimate partner violence (IPV) | 16.8 | 16.4-17.2 | 18.2 | 16.7-19.8 | 8.2 | 7.5-8.9 |
| Substance abuse in household | 26.8 | 26.3-27.3 | 29.6 | 27.8-31.5 | 15.0 | 14.1-16.0 |
| Mental illness in household | 18.4 | 17.9-18.8 | 31.5 | 29.6-33.4 | 5.4 | 4.8-6.0 |
| Parental separation or divorce | 29.3 | 28.8-29.8 | 40.2 | 38.2-42.2 | 12.2 | 11.4-13.1 |
| Incarcerated household member | 8.7 | 8.4-9.1 | 15.9 | 14.6-17.4 | 1.7 | 1.4-2.0 |

<sup>a</sup> AL, DE, FL, IN, IA, MI, MS, MO, NM, ND, PA, RI, SC, TN, VA, WV, WI, OH, KS, OK, NY

Results were confirmed in logistic regression when controlled for the factors noted (Table 4), with adjusted odds ratios (AOR) increasing with increasing age for CVD and decreasing with older age for SCI. If age is added as 7 groups as in the unadjusted results shown in Table 2, that same dip at ages 65-74 is seen. Highest AOR for SCI was 4.7 for depression, followed by all 8 ACEs at 4.0. With the composite ACEs entered as a stepwise measure up to 8, and each step compared with 0 (Table 4), AORs ranged from 1.27 for 1 ACE to 4.0 for all 8. For CVD risk factors, AORs for SCI ranged from not significant for 1 risk factor to 1.76 for 5 and then dropped to 1.66 for all 6. With CVD as outcome, highest AOR was for all 6 CVD risk factors at 9.1, ranging stepwise from 1.7 for one risk factor, and for ACEs, results below 7 ACEs all had AOR<2.0 and most had P value > 0.05 while highest was 2.4 for all 8 ACEs. Notably each outcome had an AOR of 1.3 or 1.4 in the model for the other outcome measure. For SCI, alternate results in Table 4 show each added ACE (1-8) had AOR of 1.20 (1.17-1.23) and each added CVD risk factor (1-6) had AOR =1.10 (1.06-1.14). For CVD, each added CVD risk factor had an AOR of 1.42 (1.38-1.46) and each added ACE was 1.06 (1.03-1.08).

**Table 4.**
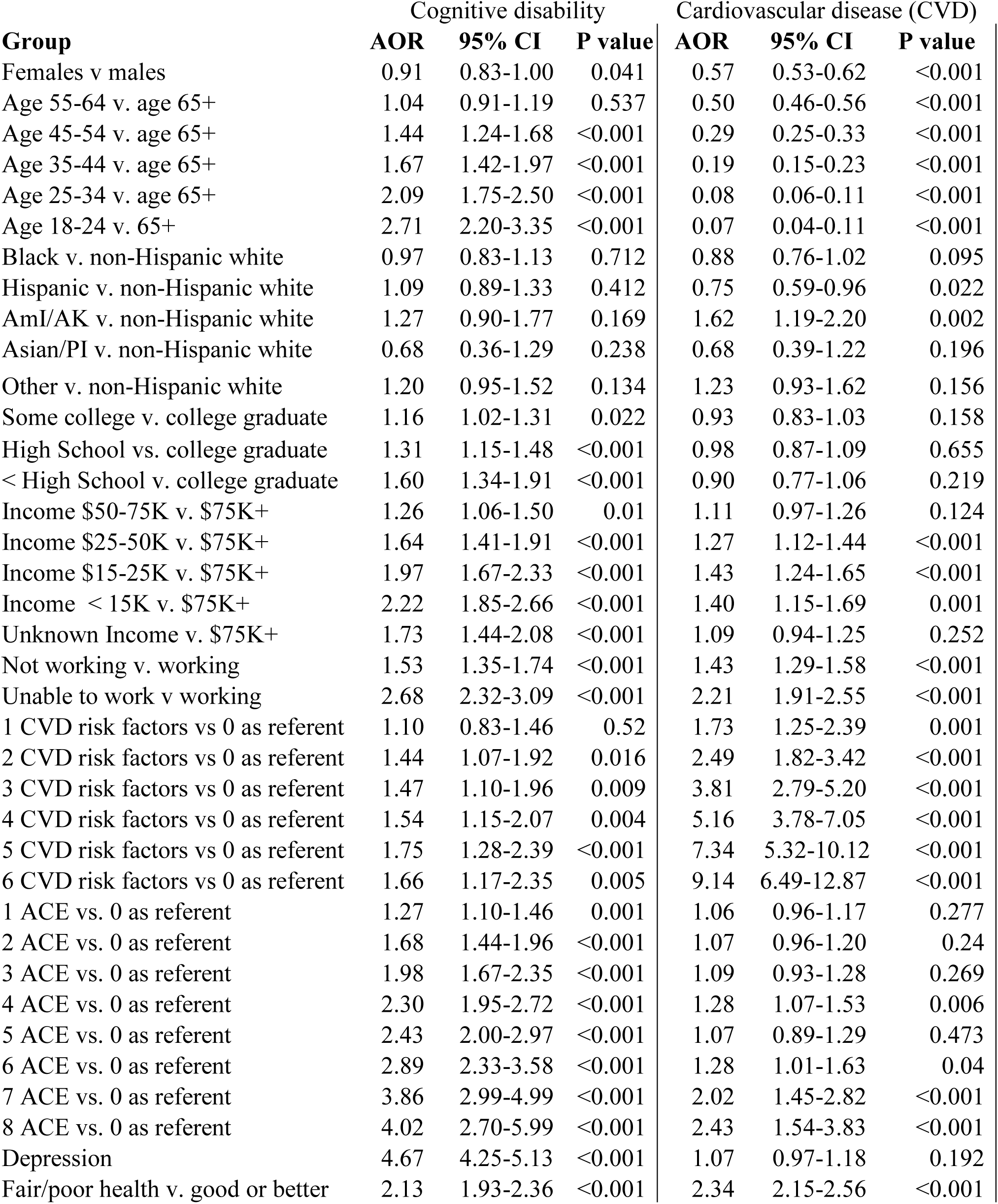
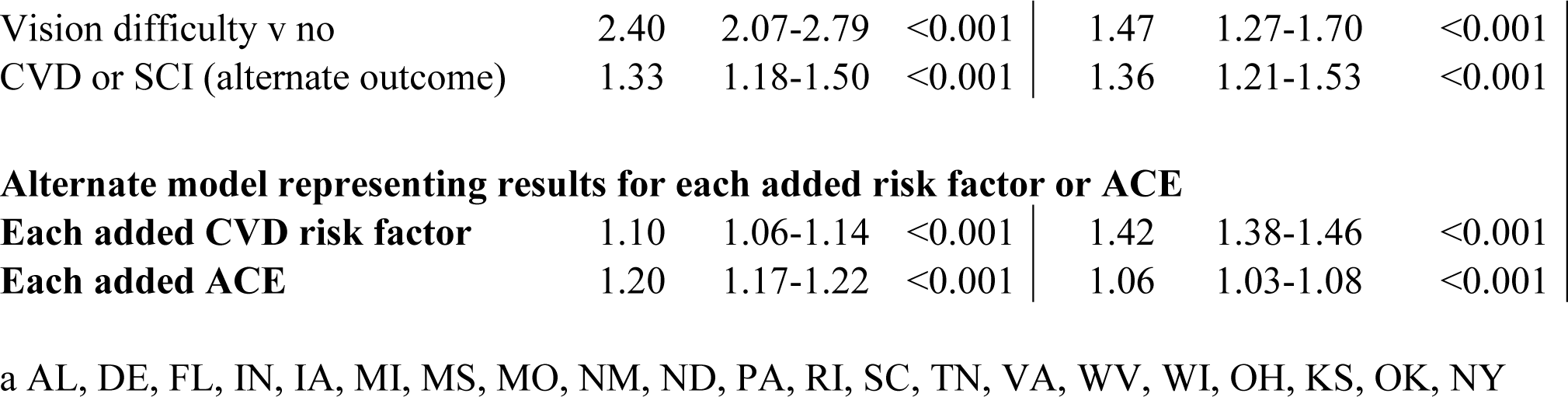
Multiple logistic regression results for Subjective Cognitive Impairment (SCI) and Cardiovascular Disease (CVD), 2019 Behavioral Risk Factor Surveillance System 21 states, N=98,325^a^.

| Group | Cognitive disability |  |  | Cardiovascular disease (CVD) |  |  |
| --- | --- | --- | --- | --- | --- | --- |
|  | AOR | 95% CI | P value | AOR | 95% CI | P value |
| Females v males | 0.91 | 0.83-1.00 | 0.041 | 0.57 | 0.53-0.62 | <0.001 |
| Age 55-64 v. age 65+ | 1.04 | 0.91-1.19 | 0.537 | 0.50 | 0.46-0.56 | <0.001 |
| Age 45-54 v. age 65+ | 1.44 | 1.24-1.68 | <0.001 | 0.29 | 0.25-0.33 | <0.001 |
| Age 35-44 v. age 65+ | 1.67 | 1.42-1.97 | <0.001 | 0.19 | 0.15-0.23 | <0.001 |
| Age 25-34 v. age 65+ | 2.09 | 1.75-2.50 | <0.001 | 0.08 | 0.06-0.11 | <0.001 |
| Age 18-24 v. 65+ | 2.71 | 2.20-3.35 | <0.001 | 0.07 | 0.04-0.11 | <0.001 |
| Black v. non-Hispanic white | 0.97 | 0.83-1.13 | 0.712 | 0.88 | 0.76-1.02 | 0.095 |
| Hispanic v. non-Hispanic white | 1.09 | 0.89-1.33 | 0.412 | 0.75 | 0.59-0.96 | 0.022 |
| AmI/AK v. non-Hispanic white | 1.27 | 0.90-1.77 | 0.169 | 1.62 | 1.19-2.20 | 0.002 |
| Asian/PI v. non-Hispanic white | 0.68 | 0.36-1.29 | 0.238 | 0.68 | 0.39-1.22 | 0.196 |
| Other v. non-Hispanic white | 1.20 | 0.95-1.52 | 0.134 | 1.23 | 0.93-1.62 | 0.156 |
| Some college v. college graduate | 1.16 | 1.02-1.31 | 0.022 | 0.93 | 0.83-1.03 | 0.158 |
| High School vs. college graduate | 1.31 | 1.15-1.48 | <0.001 | 0.98 | 0.87-1.09 | 0.655 |
| < High School v. college graduate | 1.60 | 1.34-1.91 | <0.001 | 0.90 | 0.77-1.06 | 0.219 |
| Income \$50-75K v. \$75K+ | 1.26 | 1.06-1.50 | 0.01 | 1.11 | 0.97-1.26 | 0.124 |
| Income \$25-50K v. \$75K+ | 1.64 | 1.41-1.91 | <0.001 | 1.27 | 1.12-1.44 | <0.001 |
| Income \$15-25K v. \$75K+ | 1.97 | 1.67-2.33 | <0.001 | 1.43 | 1.24-1.65 | <0.001 |
| Income < 15K v. \$75K+ | 2.22 | 1.85-2.66 | <0.001 | 1.40 | 1.15-1.69 | 0.001 |
| Unknown Income v. \$75K+ | 1.73 | 1.44-2.08 | <0.001 | 1.09 | 0.94-1.25 | 0.252 |
| Not working v. working | 1.53 | 1.35-1.74 | <0.001 | 1.43 | 1.29-1.58 | <0.001 |
| Unable to work v working | 2.68 | 2.32-3.09 | <0.001 | 2.21 | 1.91-2.55 | <0.001 |
| 1 CVD risk factors vs 0 as referent | 1.10 | 0.83-1.46 | 0.52 | 1.73 | 1.25-2.39 | 0.001 |
| 2 CVD risk factors vs 0 as referent | 1.44 | 1.07-1.92 | 0.016 | 2.49 | 1.82-3.42 | <0.001 |
| 3 CVD risk factors vs 0 as referent | 1.47 | 1.10-1.96 | 0.009 | 3.81 | 2.79-5.20 | <0.001 |
| 4 CVD risk factors vs 0 as referent | 1.54 | 1.15-2.07 | 0.004 | 5.16 | 3.78-7.05 | <0.001 |
| 5 CVD risk factors vs 0 as referent | 1.75 | 1.28-2.39 | <0.001 | 7.34 | 5.32-10.12 | <0.001 |
| 6 CVD risk factors vs 0 as referent | 1.66 | 1.17-2.35 | 0.005 | 9.14 | 6.49-12.87 | <0.001 |
| 1 ACE vs. 0 as referent | 1.27 | 1.10-1.46 | 0.001 | 1.06 | 0.96-1.17 | 0.277 |
| 2 ACE vs. 0 as referent | 1.68 | 1.44-1.96 | <0.001 | 1.07 | 0.96-1.20 | 0.24 |
| 3 ACE vs. 0 as referent | 1.98 | 1.67-2.35 | <0.001 | 1.09 | 0.93-1.28 | 0.269 |
| 4 ACE vs. 0 as referent | 2.30 | 1.95-2.72 | <0.001 | 1.28 | 1.07-1.53 | 0.006 |
| 5 ACE vs. 0 as referent | 2.43 | 2.00-2.97 | <0.001 | 1.07 | 0.89-1.29 | 0.473 |
| 6 ACE vs. 0 as referent | 2.89 | 2.33-3.58 | <0.001 | 1.28 | 1.01-1.63 | 0.04 |
| 7 ACE vs. 0 as referent | 3.86 | 2.99-4.99 | <0.001 | 2.02 | 1.45-2.82 | <0.001 |
| 8 ACE vs. 0 as referent | 4.02 | 2.70-5.99 | <0.001 | 2.43 | 1.54-3.83 | <0.001 |
| Depression | 4.67 | 4.25-5.13 | <0.001 | 1.07 | 0.97-1.18 | 0.192 |
| Fair/poor health v. good or better | 2.13 | 1.93-2.36 | <0.001 | 2.34 | 2.15-2.56 | <0.001 |
| Vision difficulty v no | 2.40 | 2.07-2.79 | <0.001 | 1.47 | 1.27-1.70 | <0.001 |
| CVD or SCI (alternate outcome) | 1.33 | 1.18-1.50 | <0.001 | 1.36 | 1.21-1.53 | <0.001 |
| **Alternate model representing results for each added risk factor or ACE** |  |  |  |  |  |  |
| **Each added CVD risk factor** | 1.10 | 1.06-1.14 | <0.001 | 1.42 | 1.38-1.46 | <0.001 |
| **Each added ACE** | 1.20 | 1.17-1.22 | <0.001 | 1.06 | 1.03-1.08 | <0.001 |

a AL, DE, FL, IN, IA, MI, MS, MO, NM, ND, PA, RI, SC, TN, VA, WV, WI, OH, KS, OK, NY

Not shown are unadjusted results for separate ACEs for the two outcomes. Each separate ACE significantly increased the rate of SCI, often close to doubling the rate. Only physical or sexual abuse as child or being in a household with IPV or substance abuse significantly increased unadjusted rates of CVD, while emotional abuse, mental illness in household, and parental separation appeared to reduce rates of CVD and there was no association with incarceration. For SCI, highest AOR for a separate ACE was ∼1.5 for mental illness in the household or sexual abuse as child, followed by physical or emotional abuse as child at ∼1.3 and substance abuse in household at ∼ 1.2. For CVD, only having an incarcerated person in the household and sexual abuse as a child had significant AORs of ∼1.2.

## Discussion

This study on subjective cognitive disability adds to existing evidence that ACEs can have potential effects on cognitive function in later life.^6–8^ The magnitude of the effect of ACEs on SCI appears somewhat higher than that due to 6 CVD risk factors, finding the AOR for each of eight ACEs entered as a continuous variable was 1.20 compared to an AOR for each of 6 risk factors of 1.10. Our results also confirmed earlier results that showed dose-response gradients between CVD risk factors and SCI ^14–15^ and found even larger dose response gradients for ACEs and SCI; all these results remained even when controlled for CVD. What is very different between CVD and SCI is the association with age; for CVD, rates increase dramatically with increasing age, while for SCI, adults ages 18-24 have highest rates which decrease more gradually with increasing age. In addition, while adults reporting SCI tended to be younger than those with CVD, they were more likely to be unable to work, in fair or poor health, and report poor vision compared with adults with CVD,

Various studies of cognitive impairment or dementia have included estimates of population attributable risk (PAR) or the proportion of the outcome caused by potentially modifiable risk factors. One study using this same measure of SCI ^14^ estimated the combined attributable risk for these 6 CVD risk factors plus inadequate fruit and vegetable consumption was approximately 50%. Another study ^20^ estimated that half of all Alzheimer’s disease cases were attributable to 5 of the 6 CVD risk factors used in this current study (excluding high cholesterol), plus depression and low educational attainment. In addition to being a recognized risk factor for dementia, depression is also known to be increased among adults with ACEs.^3–5^ One study estimated that 44.1% of overall attributable risk for depression was contributed by ACEs.^4^ Thus, not only do ACEs increase the likelihood of both depression^4^ and SCI but depression was found in this study to increase the risk of SCI and in another study was shown to increase rates of dementia.^20^ Although we did not include depression as a risk factor for SCI in our study, we did control for it in our logistic regression model. Depression had the highest AOR (4.7) of any variable in the model for SCI when ACEs and risk factors were both included yet the associations of ACEs and CVD risk factors were still substantial. These study results add support to the role of both CVD risk factors and ACEs as contributing to attributable risk for both CVD and cognitive impairment.

Our results for the prevalence of ACEs are in general agreement with previous studies of BRFSS data that used different states and years.^1, 4^ For example, our study finds higher numbers of ACEs among younger adults, women,^1, 4^ and American Indian/Alaska Natives.^4^ Comparison of ACEs results in this study with earlier results using the same BRFSS questions – but different states -- suggest an increase in ACEs between 2011-14, 2015-17 and our 2019 results. For example, 2001-14 results ^1^ report a mean number of ACEs of 1.57 for all ages and 0.87 for adults ages 65+ while our comparable results are 1.71 (1.69-1.73) and 1.04 (1.01-1.06), respectively. For 2015-2017, 39.0% of all adults and 52.1% among adults ages 65+ reported zero ACEs ^4^ compared with our 2019 rates of 35.5 and 48.7, respectively. It is unclear if the apparent decrease in the rate of reporting zero ACEs and increase in the mean number of ACEs compared with earlier years represents a meaningful increase in ACEs over that time-period but it is consistent with results ^1, 4^ showing lower rates of ACES among adults ages 65 and older. Fewer ACEs reported by older adults could indicate difficulty recalling incidents farther in the past, but the results for separate ACEs suggest that might not be the case. The age associations for the separate ACEs (Table 3) are quite different but the results for incarceration seem to agree with actual incarceration trends. ^21^ The relatively slow rise in reported ACEs over time among adults aged 65+ suggests at least some ACEs effects on younger adults may be reversible.

Higher rates of SCI in younger adults coupled with these results suggest ACEs as a possible reason but that is only one possible explanation. Another potential explanation for declining rates of SCI, cognitive decline (CD), and dementia with increasing age from telephone survey data comes from a study that included proxy responses for nonrespondents on the BRFSS. ^22^ Compared with respondents with CD, non-respondents with CD (in different households) were significantly more likely to need help, report getting a dementia diagnosis and treatment. A key difference between respondents and non-respondents was that only respondents needed to be able to respond to a telephone survey. The author concluded that as cognitive decline worsened with age, adults would become unable to respond to such a survey and that to accurately represent ALL household adults, non-respondents would need to be included. The magnitude of the effect of under-reporting due to this bias was estimated to be as high as 70%. ^22^

Our study results find that the AORs of each added ACE and CVD risk factors on SCI are similar, ranging from 1.1-1.2. This suggests that PARs for each might be of the same order of magnitude and represent a sizeable fraction of attributable risk. For adults with ACEs, they cannot change the past, but they still have a chance to change behaviors that could reduce the number of CVD risk factors and thus their risk of SCI and CVD.

Study limitations include the usual due to a cross-sectional survey not being able to show cause and effect, with the added limitation that not all states were included so generalizability is unknown. ACEs could be underestimated due to recall and social undesirability that may vary by age. Other childhood experiences not addressed on this module could influence SCI. Only non-institutionalized adults were surveyed so adults in long-term care who may be more likely to have SCI are excluded. As noted above, household adults who are physically or mentally unable to respond to a survey are also excluded, which may omit potential respondents with severe cognitive problems. This exclusion could be responsible for serious underestimates of some measures.^22^ Results from this study – and any study using data on cognitive impairment or decline from the BRFSS – should clarify that results only pertain to adults who are able to respond to a telephone survey.

Despite limitations, these results are still useful and seem to suggest the potential for disturbing future trends. First, if the association between ACEs and SCI is causal, the magnitude appears similar to that for CVD risk factors which have been estimated to be attributable to about half of Alzheimer’s disease ^20^ or SCI ^14^ cases. Next, the prevalence of ACEs among children may be increasing over time as shown by results from different years and for different age groups.^1, 4^ The time trends for CVD risk factors are mixed: Smoking rates are declining^23–24^ but obesity rates, which affect most other risk factors, are increasing.^24^ The overall result of those trends could predict increased SCI and dementia rates over time. The outlook for the future is also likely to be affected by the COVID pandemic in at least two ways. Because COVID appears to affect the brain, there is already evidence suggesting that patients who recover from COVID may be at higher risk of developing cognitive decline in the future.^25–26^ Evidence from studies on the mental health of children during the pandemic suggests that such stress might be considered another adverse childhood experience.^27^ Combine all this evidence and it appears that adults, especially those who have recovered from COVID and/or had adverse events as children, will require close monitoring for cognitive impairment now and into the future. Also, whether SCI worsens over time or not, the finding that 13.5% - 17.2% of adults aged 18-34 years are reporting cognitive impairment and those with SCI often report poor health and being unable to work suggests it is already a problem. Dialogs with primary care providers for adults with SCI at any age could potentially lead to earlier diagnosis and treatment resulting in long-term benefits.^28^ This also applies to ACEs, where early assessment and intervention and more research on best methods for improving outcomes could have long lasting impacts. These results also suggest the need for studies on cognitive impairment or decline to include all adults and not limit studies to older adults such as those ages 45 and older. Consideration might also be given to again including proxy responses for the BRFSS Cognitive Decline module.^22^

## Conclusion

Study results support earlier findings showing an association between ACEs and poorer cognition and that the impact of 8 ACEs is similar to that for 6 CVD risk factors. This study confirms other results^13^ indicating that adults 18-24 years have highest rates of SCI and suggests that ACEs may be at least partially responsible for this finding. Compared with adults with CVD, adults with SCI are more likely to be unable to work, in poor health, and have vision problems while they also tend to be younger. To date, these higher SCI rates for younger adults have not translated over time to increased rates for older adults - which is good news - but suggests the need for more study. Considering higher rates of SCI and more ACEs among younger adults plus potential COVID effects could predict increased rates of cognitive issues and dementia in the future.

## Data Availability

All source data are available at: https://www.cdc.gov/brfss/data_documentation/index.htm

## Notes

### Competing Interest Statement

The authors have declared no competing interest.

### Funding Statement

This study did not receive any funding

### Author Declarations

All data are available at: https://www.cdc.gov/brfss/data_documentation/index.htm

